# Leveraging seasonal dynamics to identify the strength of disease transmission along multiple environmental pathways

**DOI:** 10.1101/2025.02.20.25322587

**Authors:** Kayoko Shioda, Miwa Watanabe, Matthew Freeman, Karen Levy, Andrew Brouwer

## Abstract

**Background:** Many pathogens spread through multiple transmission pathways, making it difficult to identify the dominant transmission pathway for a given pathogen. Including a comprehensive set of relevant pathways in infectious disease transmission (IDT) models can provide insight into the contribution of each pathway to infections and allow for comparisons of the potential impact of control strategies targeting various pathways. Seasonal patterns in pathogen prevalence, combined with environmental testing, may allow us to differentiate the contributions of different pathways.

**Methods:** We conducted an identifiability analysis for an IDT model with three transmission pathways: direct person-to-person and two indirect, environmental pathways (water-to-person and food-to-person). We ran a series of simulations to understand the conditions under which we can successfully identify the dominant transmission pathway. Specifically, we explored the effects of different magnitudes of the transmission rate for each pathway and the seasonal timing of peak pathogen concentrations in water and food.

**Results:** Under the proposed data collection approach, we were able to successfully determine the dominant transmission pathway when simulated peak seasonality in water and food contamination differed by at least one month, even when the difference in the true transmission rates was small. However, we did not always correctly estimate the transmission rates used to generate the simulated data, even if we correctly determined the relative strengths of the pathways. When the simulated contamination in water and food peaked at the same time, the IDT model failed to determine the dominant pathway, regardless of the relative magnitudes of the true transmission rates.

**Discussion:** Our analysis found that IDT models can be a powerful tool to differentiate the contribution of different pathways to human infection with relevant empirical data on pathogen contamination in exposure sources. Our findings can help researchers design future studies and determine the feasibility of a study based on the seasonality in environmental samples if such data are available.

## 1. Introduction

Enteric pathogens, such as *Campylobacter* and *Salmonella*, can spread through diverse transmission routes, including food, water, and direct contact with infected humans and animals. These multiple transmission routes complicate efforts to understand the dynamics of pathogen spread and to identify effective control measures. While progress has been made in identifying the etiology of infections, significant gaps remain in quantifying the dominant transmission pathways of each pathogen. For instance, large multi-country studies, such as the Global Enteric Multicenter Study (GEMS) [1] and the MAL-ED Study [2], have contributed valuable knowledge about the etiology of diarrhea and how it varies across locations, but specific transmission pathways were not investigated or determined. The World Health Organization (WHO) Foodborne Disease Burden Epidemiology Reference Group (FERG) provides estimates for the contribution of enteric pathogen transmission via food, animal contact, human contact, water, soil, and other sources for various foodborne pathogens by regions of the world, but these estimates rely heavily on expert opinion rather than empirical evidence and have wide uncertainty intervals [3].

The comparative exposure assessment approach has been used to determine the relative contribution of specific infection sources and identify the important sources [4]. For example, a previous study used this approach to investigate 31 potential sources of *Campylobacter* infection, such as food items (e.g., raw milk, chicken meat), animal contact (e.g., dogs, chickens, cattle, pigs), and water sources (e.g., rivers, pools, unheated tap water) [5]. The study suggested that the consumption of raw food and direct contact with animals were significant sources of *Campylobacter* infections in the Netherlands. However, these approaches cannot account for the complex interplay between different transmission pathways, such as how changes in one pathway might affect infections from other pathways by altering the prevalence of infection, the proportion of susceptible individuals, and environmental contamination. Understanding this interplay is essential for estimating the potential impact of interventions that could have cascading effects across multiple pathways [6]. Additionally, seasonal variations in the dominant pathways have not been well quantified in the literature, and understanding these variations will be key to designing effective seasonal interventions.

Infectious disease transmission (IDT) models are powerful tools to address these challenges. They can help us understand pathogen transmission across multiple pathways [6–8] and how it may vary across seasons and weather conditions [9]. When fully calibrated by appropriate empirical data, IDT models can quantify the contributions of each transmission route and track how these contributions shift across seasons. Furthermore, IDT models can simulate targeted interventions, accounting for the complex interplay between pathways, and help us compare their potential effectiveness. The results from such models can inform public health strategies and the design of effective interventions.

Despite the potential of IDT models, there are many barriers to fully calibrating IDT models with multiple pathways, particularly because the source of each individual infection is often unknown. Whole genome sequencing (WGS) can be used to pinpoint infection sources, such as contaminated food items [10], but these techniques require resources that may not be available in low- and middle-income countries (LMICs) [11,12]. WGS may also be less effective in high-transmission settings due to high variations in the pathogen genome [13]. A potential alternative, adopting a more population-level perspective, is to collect data on human infections in parallel with pathogen concentration data in various potential sources (e.g., food items, drinking water) over time and incorporate them in IDT models. For example, a field study can be designed to recruit diarrheal cases at clinics or through household visits and simultaneously collect samples of potential infection sources (e.g., food and water) from households or communities over multiple seasons or years. Such longitudinal environmental surveillance data could help IDT models identify the dominant transmission pathways and how they vary across seasons.

Therefore, the goal of this study was to investigate when such population-level alternative approaches are effective at calibrating a multi-pathway IDT model. Specifically, we sought to address questions such as: Can an IDT model identify the dominant pathway if pathogen concentrations in various infection sources (e.g., water and food) peak around the same time? What happens if two infection sources (e.g., food and water) exhibit opposite seasonality, with one peaking in the wet season and the other in the dry season? To explore these scenarios, we conducted identifiability analyses to determine the conditions necessary for IDT models to reliably estimate transmission rates across multiple pathways. We developed an IDT model that reflects characteristics of pathogens that spread directly from infected individuals or indirectly from environmental sources, such as *Campylobacter* and *Salmonella*. We investigated the capabilities and limitations of IDT models in disentangling complex transmission dynamics across multiple pathways by using simulated human infection prevalence data—where the specific transmission pathway for each infection is unknown to reflect the real-world situation—and simulated pathogen contamination data in infection sources. Findings from our study may inform future study designs and sampling strategies for researchers interested in understanding infection spread across multiple pathways.

## 2. Methods

### 2.1 Study overview

The goal of this study was to conduct the identifiability analysis and assess under which conditions the IDT model can 1) correctly determine the dominant transmission pathway and 2) correctly determine each pathway’s transmission rate based on longitudinal environmental surveillance data and human infection data. To reflect the characteristics of pathogens that spread through multiple pathways, we developed an IDT model with the following three transmission pathways: direct person-to-person pathway and two indirect environmental pathways influenced by external forcing functions. These environmental pathways could represent any source of infection for which concentration data can be collected, such as food, water, animals, and soil. In this study, we referred to them as water-to-person and food-to-person pathways for simplicity, as they are common for enteric pathogens such as *Campylobacter* and *Salmonella* [3]. Our simulation aimed to reflect high-transmission settings where indirect environmental transmission is a significant concern, which is particularly common in LMICs [3,14,15].

We simulated the prevalence of human infection, using various combinations of predefined “true” values of the pathway-specific transmission rates and varying seasonality in food and water contamination. To estimate the transmission rates, the IDT model was fit to the simulated human prevalence data, without prior knowledge of the true source of each infection, as is often the case in real-world scenarios. In our hypothetical scenarios, the simulated field study tested 50 human samples per day and measured contamination levels in water and food on a daily basis for two years.

### 2.2 IDT model structure

We developed an IDT model using a susceptible-infectious-recovered-susceptible compartmental modeling framework, which explicitly incorporates transmission through contaminated water and food. The IDT model simulates the dynamic fractions of individuals who are susceptible to infection (*S*(*t*)), infected through either person-to-person transmission (*I*_*hh*_*(t)*), water-to-person transmission (*I*_*hw*_*(t)*), or food-to-person transmission (*I*_*hf*_*(t)*), and those who have recovered from infection (*R(t)*) on day *t* (Figure 1). The total prevalence of infections through all pathways on day *t* (*I(t)*) was calculated by adding the fraction of infections through each pathway (*I*_*hh*_*(t)* + *I*_*hw*_*(t)* + *I*_*hf*_*(t)*). Below are the ordinary differential equations describing this IDT model (Eq. 1-5):

**Figure 1.**
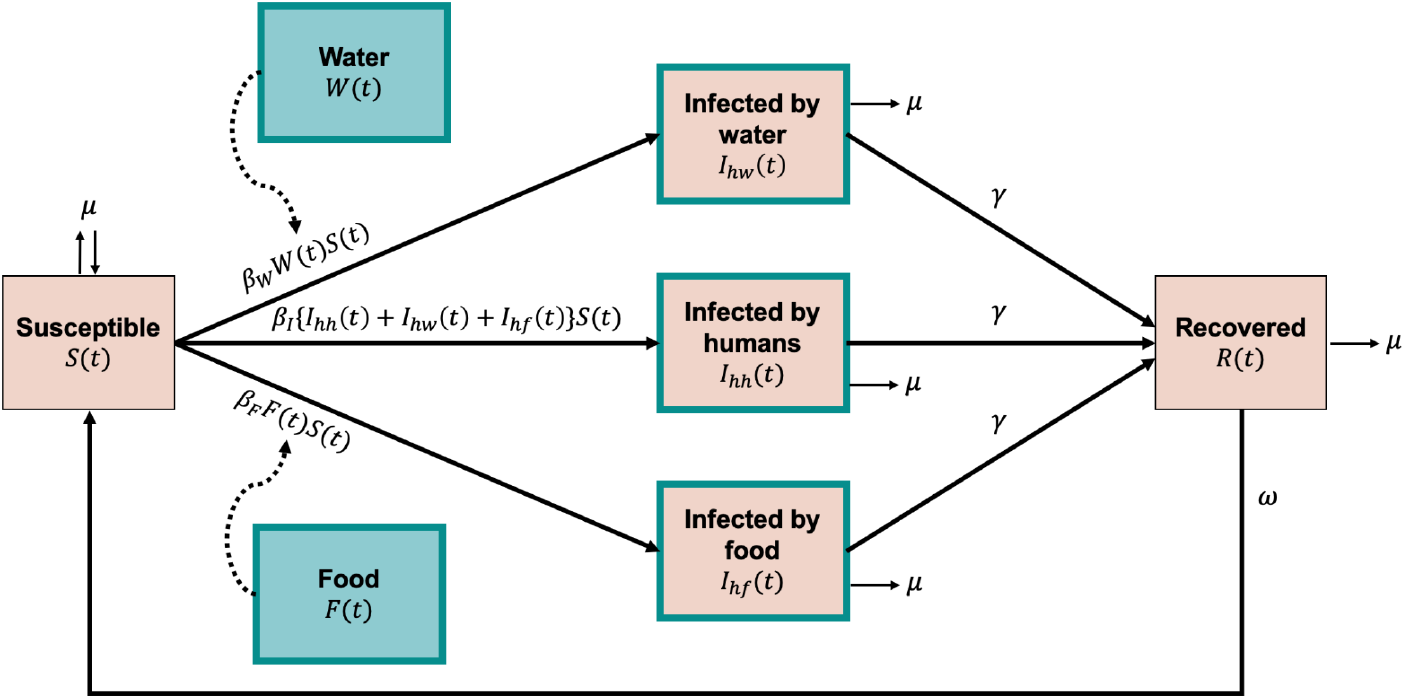
Structure of the infectious disease transmission model with multiple transmission pathways. Definitions of compartments and parameters can be found in the text and tables, but briefly:

- *S*(*t*) – the fraction of individuals who are susceptible to infection at time *t*
- *I*_*hh*_(*t*) – the fraction of individuals who are infected through person-to-person transmission at time *t*
- *I*_*hw*_(*t*) – the fraction of individuals who are infected through water-to-person transmission at time *t*
- *I*_*hf*_(*t*) – the fraction of individuals who are infected through food-to-person transmission at time *t*
- *R*(*t*) – the fraction of individuals who have recovered from infection at time *t*
- *F*(*t*) – pathogen concentration in food at time *t*
- *W*(*t*) – pathogen concentration in water at time *t*
- *β*_*I*_ – rate of transmission through person-to-person
- *β*_*W*_ – rate of transmission from water
- *β*_*F*_ – rate of transmission from food
- *μ* – birth rate and mortality rate
- *γ* – rate of recovery
- *ω* – rate of waning immunity

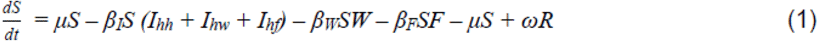

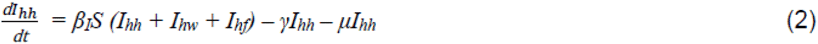

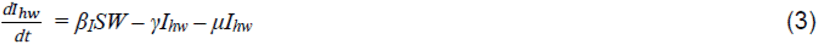

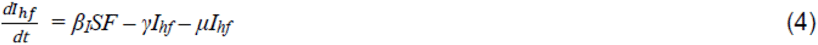

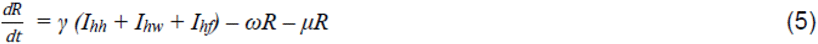

In Eq. 1-5, μ represents both the birth and mortality rates, resulting in a stable population size over time. Transmission rates for each pathway are denoted as follows: *β*_*I*_ for person-to-person, *β*_*w*_ for water-to-person, and *β*_*F*_ for food-to-person transmission. Individuals recover from infection at a rate of γ, and their immunity wanes at a rate of *ω*. Values of *μ*, *γ*, and *ω* were fixed as listed in Table 1, while the transmission rates (*β*_*I*_, *β*_*W*_, and *β*_*F*_) were estimated as described in Section 2.6. The fixed recovery (γ) and waning immunity (*ω*) rates were chosen to reflect the general characteristics of common enteric pathogens [16–19]. The birth and mortality rates were chosen to reflect those of Mozambique, as an example of an LMIC [20]. Pathogen contamination in water and food is represented by *W(t)* and *F(t)*, respectively, and was simulated as described in Section 2.4 and Eq. 6-7.

**Table 1.**
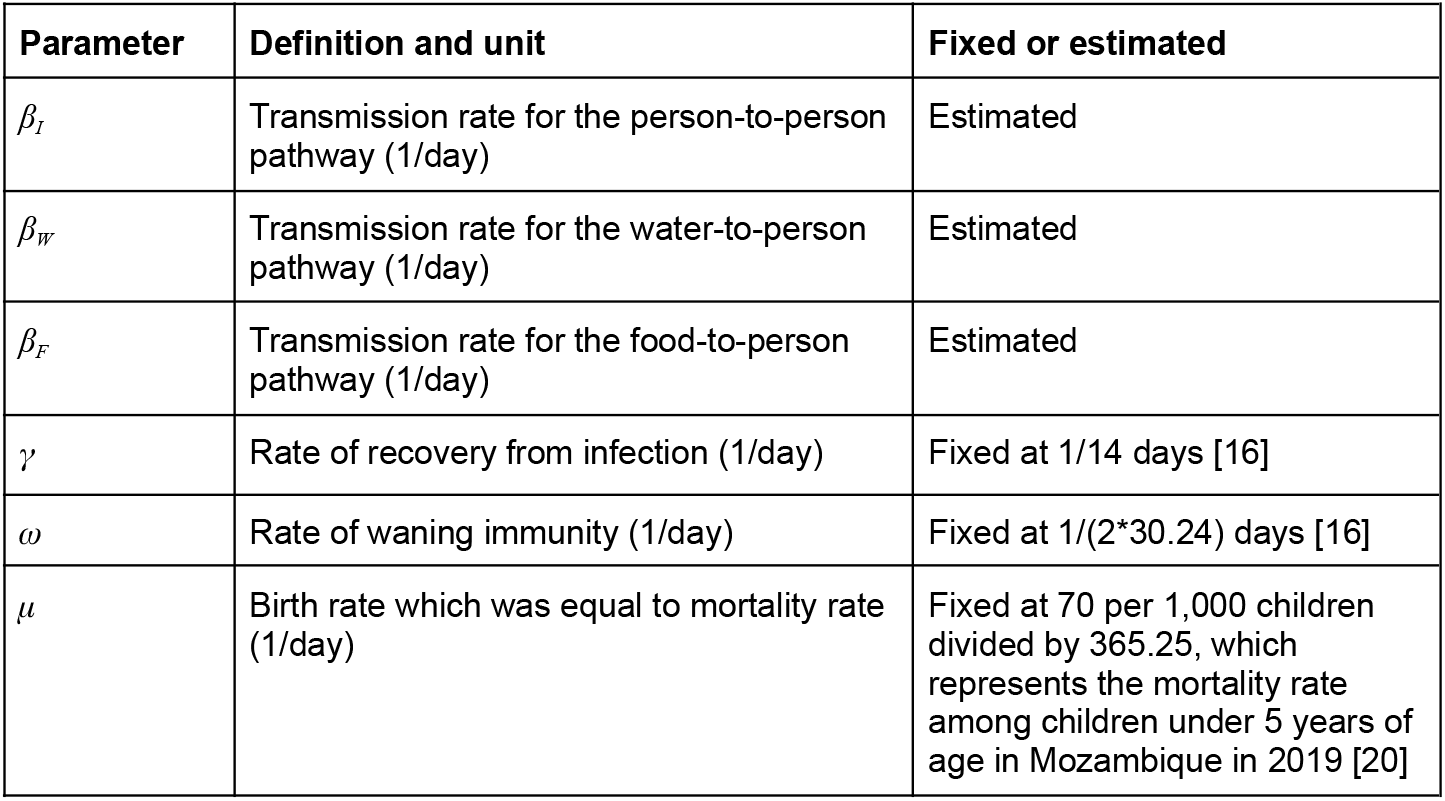
Parameters for the transmission model with multiple transmission routes.

| Parameter | Definition and unit | Fixed or estimated |
| --- | --- | --- |
| $\beta_I$ | Transmission rate for the person-to-person pathway (1/day) | Estimated |
| $\beta_W$ | Transmission rate for the water-to-person pathway (1/day) | Estimated |
| $\beta_F$ | Transmission rate for the food-to-person pathway (1/day) | Estimated |
| $\gamma$ | Rate of recovery from infection (1/day) | Fixed at 1/14 days [16] |
| $\omega$ | Rate of waning immunity (1/day) | Fixed at 1/(2*30.24) days [16] |
| $\mu$ | Birth rate which was equal to mortality rate (1/day) | Fixed at 70 per 1,000 children divided by 365.25, which represents the mortality rate among children under 5 years of age in Mozambique in 2019 [20] |

### 2.3 Investigated scenarios

To investigate when the IDT model can identify the dominant pathway, we tested a total of 16 scenarios. These scenarios combine four different timings of the annual peaks in water and food contamination (Table 2) with four sets of true values of the pathway-specific transmission rates (Table 3).

**Table 2.** Investigated scenarios by the predefined seasonality in water contamination (*W(t)*) and food contamination (*F(t)*).

| Investigated scenarios | Peak timing in water contamination (value of $\phi_W$ in Eq. 6) | Peak timing in food contamination (value of $\phi_F$ in Eq. 7) |
| --- | --- | --- |
| **Scenario i** (same peak timing) | January (365/4) | January (365/4) |
| **Scenario ii** (one month apart, with $W(t)$ peaking earlier) | January (365/4) | February (365/6) |
| **Scenario iii** (three months apart, with $W(t)$ peaking earlier) | January (365/4) | April (0) |
| **Scenario iv** (six months apart) | January (365/4) | July (-365/4) |

**Table 3.**
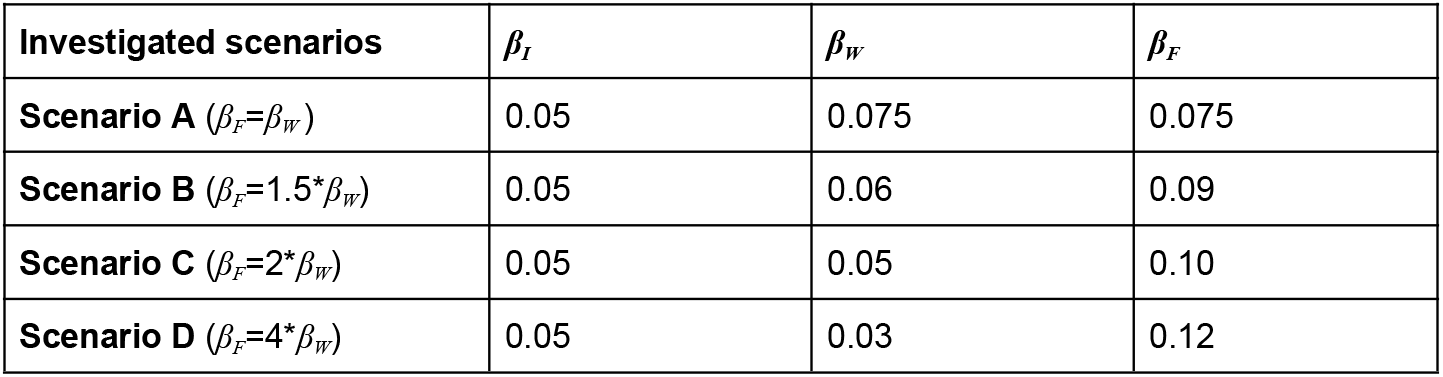
Investigated scenarios by the predefined true value of transmission rates for the person-to-person (*β*_*I*_), water-to-person (*β*_*W*_), and food-to-person (*β*_*F*_) pathways.

| Investigated scenarios | $\beta_I$ | $\beta_W$ | $\beta_F$ |
| --- | --- | --- | --- |
| **Scenario A** ($\beta_F=\beta_W$) | 0.05 | 0.075 | 0.075 |
| **Scenario B** ($\beta_F=1.5*\beta_W$) | 0.05 | 0.06 | 0.09 |
| **Scenario C** ($\beta_F=2*\beta_W$) | 0.05 | 0.05 | 0.10 |
| **Scenario D** ($\beta_F=4*\beta_W$) | 0.05 | 0.03 | 0.12 |

We modeled the seasonality of pathogen concentration in water (*W(t)*) and food (*F(t)*) using sine functions so that each had an annual peak. Four scenarios were created (Table 2): in Scenario *i*, pathogen concentration in water (*W(t)*) and food (*F(t)*) had identical peak timing (January); in Scenario *ii*, their peaks were offset by one month (*W(t)* peaking in January and *F(t)* in February); in Scenario *iii*, their peaks were offset by three months (*W(t)* peaking in January and *F(t)* in April); and in Scenario *iv*, the peaks were six months apart representing complete opposite seasonality (*W(t)* peaking in January and *F(t)* in July).

We selected the “true” values of three pathway-specific transmission rates (*β*_*I*_, *β*_*W*_, and *β*_*F*_) so that the average prevalence of human infection was approximately 15%, consistent with major zoonotic enteropathogens in low-income settings [21]. In all scenarios, *β*_*I*_ was held constant at 0.05. We set the sum of the remaining 2 transmission rates (*β*_*W*_ + *β*_*F*_) to 0.15 in all simulated scenarios (Table 3). In Scenario A, *β*_*W*_ and *β*_*F*_ were equal. In Scenario B, *β*_*F*_ was 1.5 times larger than *β*_*W*_. In Scenario C, *β*_*F*_ was twice as large as *β*_*W*_. In Scenario D, *β*_*F*_ was four times larger than *β*_*W*_.

### 2.4 Simulation of the “true” external forcing functions and its observed data

We modeled “true” daily time series of pathogen contamination data in water (*W*(t)*) and food (*F*(t)*) using sinusoidal functions as follows:

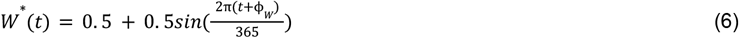

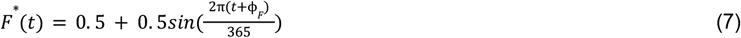

The functions above generate values within the interval [0, 1], with a mean of 0.5 (Supplementary Figure 1-A). In practice, pathogen concentration data are typically obtained by various laboratory assays, such as PCR (quantified in copies per volume) or culture-based methods (quantified in colony-forming units per volume), where observed values can range from zero to infinity. For the purpose of this simulation, we assumed that these pathogen concentration data were standardized to constrain values to the interval [0, 1], maintaining a mean of 0.5.

The phase terms, *φ*_*W*_ and *φ*_*F*_, were varied to represent different annual seasonal peak timings for water and food contamination, as detailed in Section 2.3 and Table 2. The observed daily data on water contamination (*W*_*obs*_*(t)*) and food contamination (*F*_*obs*_*(t)*) by adding noise to *W*(t)* and *F*(t)* using a normal distribution with a mean of *W*(t)* or *F*(t)* and a standard deviation of 0.25 (Supplementary Figure 1-B). To ensure all values remained within the interval [0, 1], any values falling below zero were set to zero, and any exceeding one were capped at one.

Lastly, the observed data were smoothed by fitting sine curves, resulting in 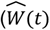 and 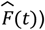, which is a common approach (Supplementary Figure 1-C). The true daily time series data (*W*(t)* and *F*(t)*) were used to simulate the true prevalence of human infection (see Section 2.5), while the smoothed observed data 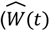 and 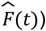 were used for parameter estimation (Section 2.6).

### 2.5 Simulation of the “true” prevalence of human infection and its observed data

The true prevalence of human infection was simulated using the IDT model detailed in Section 2.2 and Figure 1. For each investigated scenario, we ran this IDT model for three years (*t* = 1, …, 365 x 3) with parameter values in Table 1-3. The simulated data for the first year was treated as a burn-in period and not included in the parameter estimation to ensure that the model had reached the periodic dynamics.

The true daily time series of water and food contamination (*W*(t)* and *F*(t)*) were used to simulate the true prevalence of human infection (Figure 2). To simulate the observed number of infections on day *t* (*x* _*t*_ ), a binomial distribution was used to model how many of the human samples (*n*_*t*_ =50) tested positive on day *t*, where the probability was *I(t)*.

**Figure 2.**
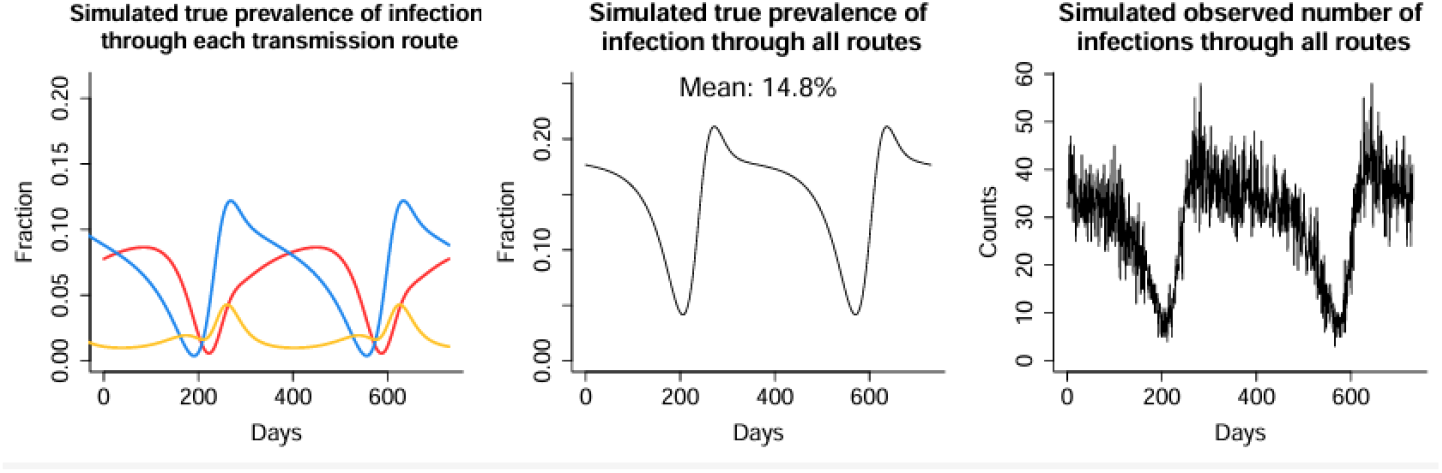
Simulated daily time series data on the true and observed prevalence of human infection (Scenario ii-A as an example). Left: Each line represents the simulated true daily fraction of the population infected through each transmission pathway (orange: person-to-person, blue: water-to-person, red: food-to-person).

### 2.6 Parameter optimization and profile likelihood analysis

We connected the model parameters to the simulated data, using the following binomial log-likelihood as our outcome (infection) was binary:

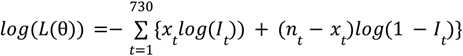

Where *L*(θ) is the likelihood and θ is a set of parameters to be estimated (*β*_*I*_, *β*_*W*_, and *β*_*F*_). The negative log-likelihood (NLL) can be defined as:

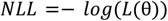

A cost function, χ^2^, can then be defined as:

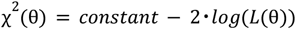

To find the maximum likelihood parameter estimates (i.e., the parameters minimizing the NLL) and construct 95% confidence intervals (CIs) for the estimated transmission rates, we performed profile likelihood analyses [22]. Profile likelihood, 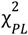, was calculated for each of the estimated parameters, θ_*i*_ , as follows [23,24]:

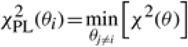

For example, to conduct profile likelihood analysis for *β*_*W*_, we first created a vector of 200 candidate values for *β*_*W*_ . For each of these candidate values, we optimized χ ^2^(θ) with respect to *β*_*I*_ and *β*_*F*_. The relative cost function was calculated by subtracting the minimum negative log-likelihood from the negative log-likelihood at each candidate value. The chi-square distribution was then used to determine the threshold, Δ_α_, for identifying the lower and upper bounds of the CIs as follows:

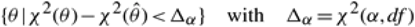

where the degree of freedom, df, was 3 and α was 0.05.

### 2.7 Evaluation of model performance

For each investigated scenario, we assessed the performance of the IDT model by examining its ability to correctly determine the predefined ordering of the rates for the water, food, and person-to-person transmission pathways. This focus reflects real-world scenarios, where the primary objective is to identify the dominant transmission pathway in each season to guide the design of effective, season-specific interventions. Additionally, we evaluated whether the predefined true values of transmission rates fell within the 95% CIs generated based on profile likelihood analysis as a secondary metric.

### 2.8 Alternative scenarios

Under all investigated scenarios, *β*_*I*_ was set to 0.05, the smallest value among all pathways in nearly all scenarios (Section 2.3 and Table 3). This choice reflects the characteristics of common enteropathogens in LMICs, where environmental transmission is predominant [3,14,15]. To assess model performance under conditions where *β*_*I*_ is the largest, we conducted an analysis with *β*_*I*_ = 0.11, *β*_*W*_ = 0.03, and *β*_*F*_ = 0.06. The sum of all *β* values remained 0.2, consistent with the main investigated scenarios.

### 2.9 Software and Code

All analyses were conducted with R version 4 (R Center for Statistical Computing), and our code can be found in the supplementary material as well as in the following GitHub repository: https://github.com/KayokoShioda/SIRS_WF_Identifiability.

## 3. Results

### 3.1 Overall results

The IDT model successfully identified the food-to-person pathway as dominant in seven out of the 12 scenarios where *β*_*F*_ was predefined to be greater than *β*_*W*_ and *β*_*I*_ (Table 4). The model reliably determined the dominant pathway when the seasonal peaks of water and food contamination differed by one and three months (Scenario *ii-vi*), even when the difference between *β*_*F*_ and *β*_*F*_ was small. However, when water and food contamination shared identical seasonal peaks (Scenario *i*), the model was unable to discern the relative magnitude of *β*_*W*_ and *β*_*F*_, regardless of how much larger *β*_*F*_ was compared to *β*_*W*_. When water and food contamination peaked six months apart (i.e., the opposite seasonality; Scenario *iv*), *β*_*I*_ was estimated to be the dominant pathway, unless the difference between the true values of *β*_*W*_ and *β*_*F*_ was large. Additionally, in many scenarios, the predefined true values were not correctly estimated (Supplementary Table 5), likely a result of error 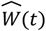 and 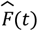 compared to *W*(t)* and *F*(t)*. For example, our data simulation approach resulted in 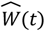 and 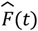 underestimating the magnitudes of the sinusoidal functions (Supplementary Figure 1), and that error propagated into the estimates of the transmission rates. Below, we summarized the model performance, evaluated based on the metric described in Section 2.7, under each investigated scenario.

**Table 4.** Results of the structural identifiability analysis across 16 investigated scenarios.

| Investigated scenarios | Scenario i<br>(same peak timing) | Scenario ii<br>(one month apart, with $W(t)$ peaking earlier) | Scenario iii<br>(three months apart, with $W(t)$ peaking earlier) | Scenario iv<br>(six months apart) |
| --- | --- | --- | --- | --- |
| **Scenario A**<br>($\beta_F = \beta_W$) | All rates not identifiable | $\beta_F < \beta_W$<br>($\beta_I$ not identifiable) | $\beta_F < \beta_W$<br>($\beta_I$ not identifiable) | $\beta_I$<br>($\beta_W$ and $\beta_F$ not identifiable) |
| **Scenario B**<br>($\beta_F = 1.5 * \beta_W$) | All rates not identifiable | $\beta_W < \beta_F$<br>($\beta_I$ not identifiable) | $\beta_W < \beta_F$<br>($\beta_I$ not identifiable) | $\beta_F < \beta_I$<br>($\beta_W$ not identifiable) |
| **Scenario C**<br>($\beta_F = 2 * \beta_W$) | All rates not identifiable | $\beta_W < \beta_F$<br>($\beta_I$ not identifiable) | $\beta_W < \beta_F$<br>($\beta_I$ not identifiable) | $\beta_F < \beta_I$<br>($\beta_W$ not identifiable) |
| **Scenario D**<br>($\beta_F = 4 * \beta_W$) | All rates not identifiable | $\beta_W < \beta_F$<br>($\beta_I$ not identifiable) | $\beta_W < \beta_F$<br>($\beta_I$ not identifiable) | $\beta_W < \beta_I < \beta_F$ |

### 3.2 *Model performance under Scenario i (identical peak timing of W(t)* and *F(t)*)

Under Scenario *i*, where *W(t)* and *F(t)* had identical seasonality (both peaking in January), the simulated true prevalence of infection from all pathways, *I(t)*, exhibited pronounced annual seasonality, ranging from 2% to 22% (Supplementary Table 1). For pathway-specific infections, *I*_*hw*_*(t)* and *I*_*hf*_*(t)* showed identical temporal patterns, with their peaks and troughs occurring simultaneously. Infections from person-to-person transmission are influenced by the prevalence of infection within the simulated population and, as a result, *I*_*hh*_*(t)* also peaked around the same time as *I*_*hw*_*(t)* and *I*_*hf*_*(t)*.

Under this scenario, where *W(t)* and *F(t)* shared identical seasonality, the IDT model was unable to uniquely estimate any of the transmission rates, regardless of their relative magnitudes (Table 4). All transmission rates could have been negligible (lower 95% CI was 0), as infections could be equally attributable to any of the pathways.

### 3.3 *Model performance under Scenario ii (peak timing of W(t)* and *F(t) offset by one month*)

In Scenario *ii*, the peak timing of *W(t)* was one month earlier than that of *F(t)* (January and February, respectively). Under this condition, *I(t)* showed strong annual seasonality, ranging from 4% to 21% (Supplementary Table 2). While *F(t)* peaked just one month after *W(t)*, the peak in *I*_*hf*_ did not come until many more months after *I*_*hw*_ (Figure 2). This is because of the proportion of susceptible individuals. Because *W(t)* peaks first, the water pathway quickly spreads infection among a large share of the susceptible population, leading to a steep slope and a tall peak in *I*_*hw*_. The smaller pool of remaining susceptibles slows the spread of infection through the food pathway, resulting in a shallower slope and a later peak in *I*_*hf*_.

When the true value of *β*_*F*_ was predefined to be larger than that of *β*_*W*_, the IDT model successfully identified the dominant pathway (Scenario *ii*-B, C, D in Table 4). When the true value of *β*_*F*_ was set to be four times larger than *β*_*W*_ (Scenario *ii*-D), *β*_*W*_ was estimated to be zero. When the true values of *β*_*W*_ and *β*_*F*_ were set to be equal (Scenario *ii*-A), the water pathway, whose contamination level peaked one month earlier, produced a higher peak in the corresponding prevalence, *I*_*hw*_*(t)*. As a result, the model estimated a slightly higher transmission rate for the water pathway; however, the difference between the estimated *β*_*W*_ and *β*_*F*_ was minimal (Supplementary Table 5).

We also ran simulations in which we reversed the peak timing of water and food contamination so that food contamination peaked one month before water contamination. The results remained the same. The model was able to determine the dominant environmental pathway across all conditions where the true value of *β*_*F*_ was larger than that of *β*_*W*_. When *β*_*W*_ and *β*_*F*_ were predefined to be the same, the model estimated that *β*_*F*_ was slightly larger than *β*_*W*_. The rate for the person-to-person transmission pathway, *β*_*I*_, was estimated to be zero under all conditions in Scenario *ii*.

### 3.4 *Model performance under Scenario iii (peak timing of W(t)* and *F(t) offset by three months*)

In Scenario *iii*, the peak timing of *W(t)* and *F(t)* was set to differ by three months. These conditions produced moderate annual seasonality in *I(t)*, with the amplitude (absolute difference between the highest and lowest prevalence) ranging from 6% to 10%, depending on the values of *β*_*W*_ and *β*_*W*_ (Supplementary Table 3).

Under Scenario *iii*, the IDT model successfully identified the dominant pathway across all conditions, even when the differences between the true values of *β*_*W*_ and *β*_*F*_ were relatively small (Table 4). These findings remained consistent regardless of whether *W(t)* and *F(t)* peaked earlier. When *β*_*W*_ and *β*_*F*_ were predefined to be equal, similar to what was observed under Scenario *ii*, the transmission pathway that peaked earlier resulted in a higher peak in the prevalence of pathway-specific infections (i.e., *I*_*hw*_*(t)* in Scenario *iii*-A) and the corresponding transmission rate was estimated to be slightly larger than that of the other pathway (Supplementary Table 5). Similar to Scenario *ii, β*_*I*_ was estimated to be zero under all conditions.

### 3.5 *Model performance under Scenario iv (opposite peak timing of W(t)* and *F(t))*

In Scenario *iv*, the peak timing of *W(t)* and *F(t)* was set six months apart, resulting in *I*_*hw*_*(t)* and *I*_*hf*_*(t)* exhibiting opposing temporal dynamics, with their respective peaks and troughs occurring six months apart. Consequently, when the true values of *β*_*W*_ and *β*_*F*_ were set to be equal (Scenario *iv*-A), *I*_*hh*_*(t)* and *I(t)* appeared flat, as the opposing seasonalities of *I*_*hw*_*(t)* and *I*_*hf*_*(t)* canceled each other out. In scenarios where *β*_*F*_ was greater than *β*_*W*_ (Scenario *iv*-B, *iv*-C, and *iv*-D), *I(t)* exhibited weak annual seasonality with a low amplitude, ranging from 1% to 5% (Supplementary Table 4).

Unlike Scenario *ii* and *iii*, the IDT model was only able to identiy the dominant pathway only when *β*_*F*_ was predefined to be four times larger than *β*_*W*_ (Scenario *iv*-D). When the difference between the true values of *β*_*W*_ and *β*_*F*_ was smaller than that (Scenario *iv*-A, B, and C), *β*_*I*_ was estimated to be the dominant pathway (Table 4). While *β*_*I*_ was estimated to be zero under Scenario *i-iii, β*_*I*_ was identifiable across all combinations of *β*_*W*_ and *β*_*F*_ values under Scenario *iv* (Supplementary Table 5).

### 3.6. Model performance under the alternative scenario

We evaluated the model performance under an additional scenario where the true value of *β*_*I*_ was set to be the largest (*β*_*I*_ = 0.11, *β*_*W*_ = 0.03, and *β*_*F*_ = 0.06), with *F(t)* and *W(t)* peaking at the same time, one, three, and six months apart. The model was able to determine the dominant environmental pathway (i.e., food) when *W(t)* and *F(t)* peaked one, three, and six months apart from each other (Supplementary Table 6). *β*_*I*_ was only identifiable when *W(t)* peaked three months before *F(t)*. When the peak timing of *W(t)* and *F(t)* was flipped (i.e., *F(t)* peaking three months before *W(t)*), *β*_*I*_ was estimated to be zero. Similarly, when there were six months between the peaks of *W(t)* and *F(t), β*_*I*_ was estimated to be zero.

## 4. Discussion

IDT models can represent the complex transmission dynamic of pathogens across multiple pathways, including direct person-to-person transmission and indirect environmental routes. By integrating longitudinal environmental surveillance data, these models can quantify the relative contribution of different transmission pathways to human infections. These models can be used to simulate potential interventions targeting different pathways, providing critical insights into comparative effectiveness, accounting for non-linear relationships of multiple pathways. Our identifiability analysis showed that the IDT model reliably identified the dominant transmission pathway when the annual seasonality of external forcing functions (e.g., pathogen contamination in water and food) differed by one or three months, even when the difference in true transmission rates was small. However, when external forcing functions shared identical seasonal patterns, the model was incapable of determining the dominant pathway, even when the true transmission rates differed considerably. Additionally, even the relatively minor noise added in our simulated data resulted in misestimation of the actual transmission rates in most scenarios, highlighting that accurate model calibration remains a challenge, even when we can correctly identify the dominant transmission pathway.

In real-world settings, it is rarely possible to trace individual infections back to their specific source, particularly in high-transmission environments. This limitation makes it difficult to quantify the prevalence of infections attributable to each transmission pathway. IDT models address this challenge by integrating human infection data with environmental data. By fitting IDT models, informed by the information on external forcing functions, to the overall prevalence of infection aggregated across all pathways, the model can estimate pathway-specific transmission rates and simulate the prevalence of infections from each pathway. This approach enables us to identify dominant transmission pathways for different seasons and design effective, seasonally tailored interventions.

Understanding how dominant pathways shift with seasons or weather conditions is increasingly important in the era of climate change. For instance, studies have shown that water sources are more heavily contaminated with fecal pathogens during the rainy season compared to the dry season [25], and contamination levels in food items also vary across seasons [26,27]. These findings underscore the need for interventions that are adapted to seasonal changes.

Additionally, understanding seasonal shifts in dominant pathways can help predict disease burden under specific climate change scenarios and guide the development of targeted mitigation plans. While existing projections of future enteric pathogen incidence under various climate scenarios provide valuable insights [28], they often overlook the distinct ways different transmission routes may respond to climate change. The framework presented here offers a tool to bridge this gap, enabling more nuanced and actionable predictions.

While our analysis referred to indirect environmental transmission pathways as water and food for simplicity, the IDT model can be adapted to investigate other pathways, such as animal contact or soil, provided longitudinal surveillance data are available. For instance, it could be used to compare the contributions of specific water sources (e.g., tap water, stored water, river water) or food items (e.g., meat, milk, grains, salads). Similarly, the model could assess the relative importance of transmission through various animal species (e.g., cattle, pigs, goats, chickens). These analyses can help us design targeted interventions based on the most significant sources.

We evaluated the model performance by assessing its ability to correctly determine the relative strengths of *β*_*W*_ and *β*_*F*_. The identifiability of each transmission rate was also evaluated as a secondary metric. In many scenarios, the 95% CIs based on the profile likelihood analysis did not include the predefined true value of each transmission rate, although many were close. The likely source of this discrepancy is the noise added to the simulated data on water contamination, food contamination, and human infections. This result highlights the real challenges for the calibration of IDT models with environmental pathways, and future work will need to investigate how robust the simulated outcomes of interventions are in the face of typical calibration error. Nevertheless, our primary evaluation metric in this analysis is relevant to real-world situations, as the main concern in practice is designing transmission control measures that target the dominant pathway across seasons, rather than obtaining accurate estimates of the transmission rates.

Our simulation analyses provided interesting findings on the non-linear relationship among multiple transmission pathways. When the peak timing of *W(t)* and *F(t)* was one or three months apart, even when the true values of *β*_*W*_ and *β*_*F*_ were predefined to be equal, the transmission pathway that peaked earlier (after the seasonal nadir in transmission) yielded a higher peak in the prevalence of pathway-specific infections, and the corresponding transmission rate was estimated to be larger than the other. This result is interesting because one might expect the prevalence of pathway-specific infections to be equal due to the same predefined values of *β*.

This difference is likely due to the prevalence of susceptible individuals on day *t*. The simulated proportion of susceptible individuals increased over the off-season when both water and food pathways had low prevalence, providing more opportunities for the transmission pathway that peaks earlier to spread quickly. For the other pathway that peaked later, there is only a smaller pool of susceptibles left, resulting in a slower spread of infection. Hence, a pathway that is “dominant” in the sense that the transmission rate is larger may not be “dominant” in the sense of attributable fraction of cases. These findings highlight the complex relationships between multiple transmission pathways and the importance of using transmission modeling to understand these non-linear relationships.

The rate for person-to-person transmission, *β*_*I*_, was estimated to be zero under many scenarios, except when *W(t)* and *F(t)* peaked 6 months apart. This result is likely because of bias in estimating the external forces (food and water contamination); the magnitudes of the estimated forcing functions were smaller than their true values, so that the constant person-to-person transmission could instead be explained by the minimum levels of the estimated external forcing functions (i.e., the estimated forcing function were never 0, so the model needed to attribute person-to-person infections to the forcing functions). In contrast, when *W(t)* and *F(t)* exhibited the opposite seasonality, the opposite issue arose, where cases caused by the external forcing functions were instead attributed to person-to-person infection. The complete opposite peak timing of water and food contamination canceled out the seasonality, generating a flat prevalence of the total infections. Because the prevalence of infections from person-to-person transmission is informed by the total prevalence of infections in the population, it also exhibited a flat prevalence. Therefore, when fitting the model to the simulated data, it was considered that the person-to-person transmission pathway explained the majority of the trends in the prevalence of total infections, and the model determined that the person-to-person pathway was dominant. While it is unlikely that environmental sources exhibit completely opposite peak timings in real-world settings, our simulation analyses may be helpful for researchers to determine whether the transmission modeling is a useful approach to identify the dominant pathway under specific conditions of interest.

The study has several limitations. While this study focused on specific scenarios designed to be straightforward and easy to interpret, real-world data may exhibit more complex patterns. For instance, the seasonality of water and food contamination may not follow a sinusoidal pattern and may follow a bump function or have multiple peaks within a year. Additionally, real-world data may show year-to-year variation due to outbreaks and susceptible depletion. Moreover, pathogens may be transmitted through more than two pathways, further complicating efforts to differentiate between them. These complexities highlight the need for studies collecting data on human infections and infection sources to address these challenges.

## 5. Conclusions

IDT models can be powerful tools to differentiate the contribution of different transmission pathways to human infections and identify the dominant pathway when integrated with relevant empirical data on pathogen contamination data from exposure sources. Our IDT model and simulation analysis can help researchers design their future field projects and determine the feasibility of a study based on the seasonality in environmental samples if such data are available from previous studies or the pilot phase of the studies.

## Data Availability

All data produced are available online at GitHub repository: https://github.com/KayokoShioda/SIRS_WF_Identifiability.

## Acknowledgment

A.B. was supported by the United States National Science Foundation (NSF) grant DMS1853032. The funder of the study had no role in study design, data collection, data analysis, data interpretation, or writing of the report.

## Web Appendix

## Supplementary Tables

**Supplementary Table 1.**
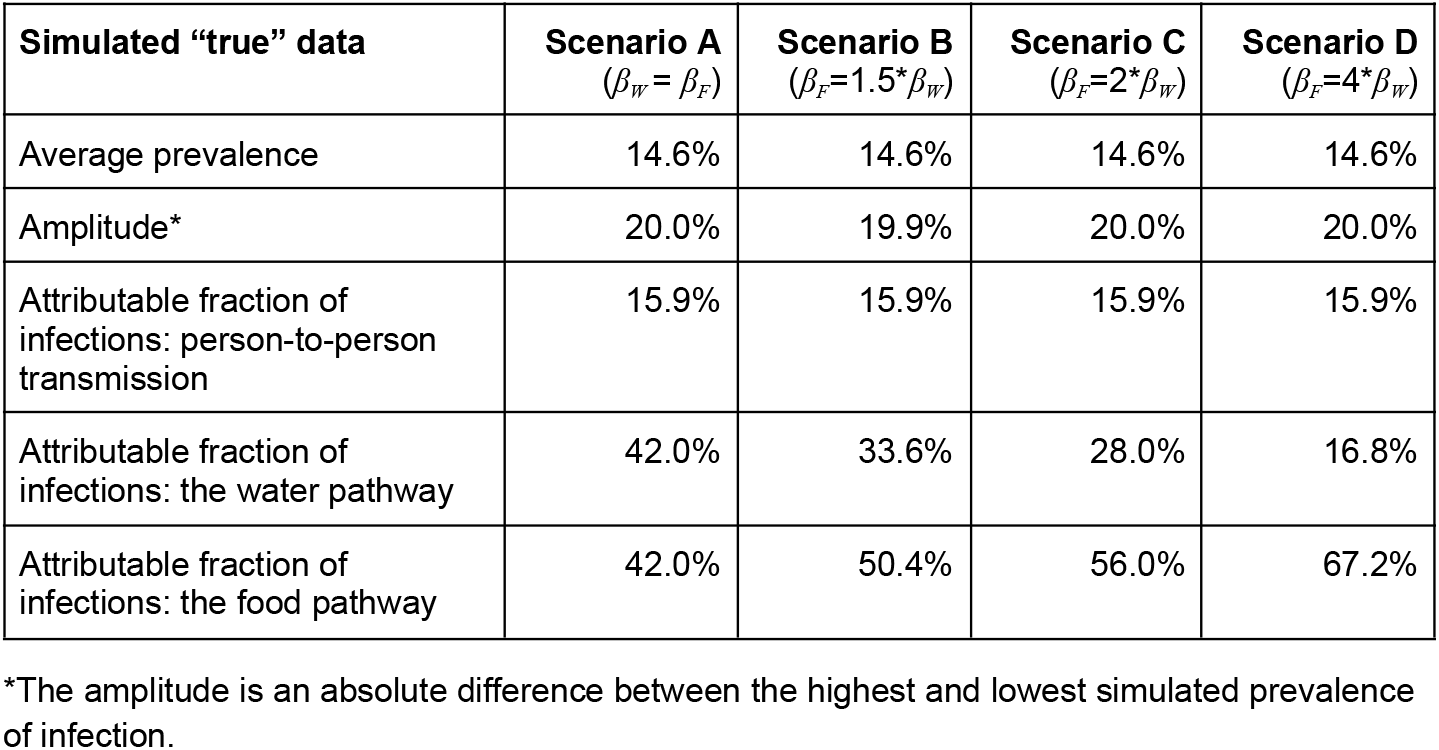
Simulated “true” prevalence of human infection and attributable fractions by transmission pathway (Scenario *i* – identical peak timing in *W(t)* and *F(t)*).

| Simulated “true” data | Scenario A<br>($\beta_W = \beta_F$) | Scenario B<br>($\beta_F = 1.5 * \beta_W$) | Scenario C<br>($\beta_F = 2 * \beta_W$) | Scenario D<br>($\beta_F = 4 * \beta_W$) |
| --- | --- | --- | --- | --- |
| Average prevalence | 14.6% | 14.6% | 14.6% | 14.6% |
| Amplitude* | 20.0% | 19.9% | 20.0% | 20.0% |
| Attributable fraction of infections: person-to-person transmission | 15.9% | 15.9% | 15.9% | 15.9% |
| Attributable fraction of infections: the water pathway | 42.0% | 33.6% | 28.0% | 16.8% |
| Attributable fraction of infections: the food pathway | 42.0% | 50.4% | 56.0% | 67.2% |

\*The amplitude is an absolute difference between the highest and lowest simulated prevalence of infection.

**Supplementary Table 2.**
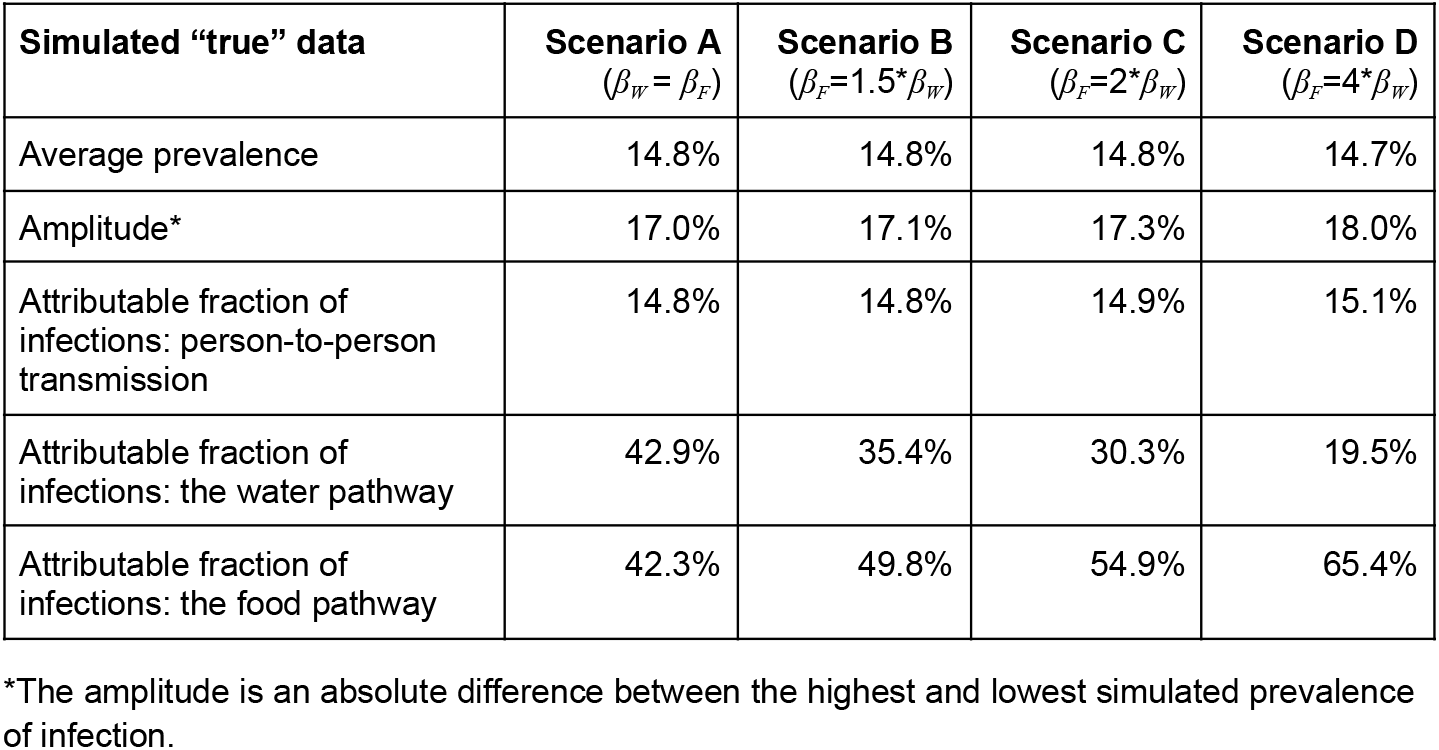
Simulated “true” prevalence of human infection and attributable fractions by transmission pathway (Scenario *ii* – *W(t)* peaks one month before *F(t)*).

| Simulated “true” data | Scenario A<br>($\beta_W = \beta_F$) | Scenario B<br>($\beta_F = 1.5 * \beta_W$) | Scenario C<br>($\beta_F = 2 * \beta_W$) | Scenario D<br>($\beta_F = 4 * \beta_W$) |
| --- | --- | --- | --- | --- |
| Average prevalence | 14.8% | 14.8% | 14.8% | 14.7% |
| Amplitude* | 17.0% | 17.1% | 17.3% | 18.0% |
| Attributable fraction of infections: person-to-person transmission | 14.8% | 14.8% | 14.9% | 15.1% |
| Attributable fraction of infections: the water pathway | 42.9% | 35.4% | 30.3% | 19.5% |
| Attributable fraction of infections: the food pathway | 42.3% | 49.8% | 54.9% | 65.4% |

\*The amplitude is an absolute difference between the highest and lowest simulated prevalence of infection.

**Supplementary Table 3.** Simulated “true” prevalence of human infection and attributable fractions by transmission pathway (Scenario *iii* – *W(t)* peaks three months before *F(t)*).

| **Simulated “true” data** | **Scenario A**<br>($\beta_W = \beta_F$) | **Scenario B**<br>($\beta_F = 1.5 * \beta_W$) | **Scenario C**<br>($\beta_F = 2 * \beta_W$) | **Scenario D**<br>($\beta_F = 4 * \beta_W$) |
| --- | --- | --- | --- | --- |
| Average prevalence | 15.7% | 15.7% | 15.6% | 15.4% |
| Amplitude* | 6.6% | 6.8% | 7.4% | 9.7% |
| Attributable fraction of infections: person-to-person transmission | 11.6% | 11.7% | 11.9% | 12.6% |
| Attributable fraction of infections: the water pathway | 44.2% | 38.4% | 34.3% | 25.1% |
| Attributable fraction of infections: the food pathway | 44.1% | 49.8% | 53.8% | 62.2% |

\*The amplitude is an absolute difference between the highest and lowest simulated prevalence of infection.

**Supplementary Table 4.**
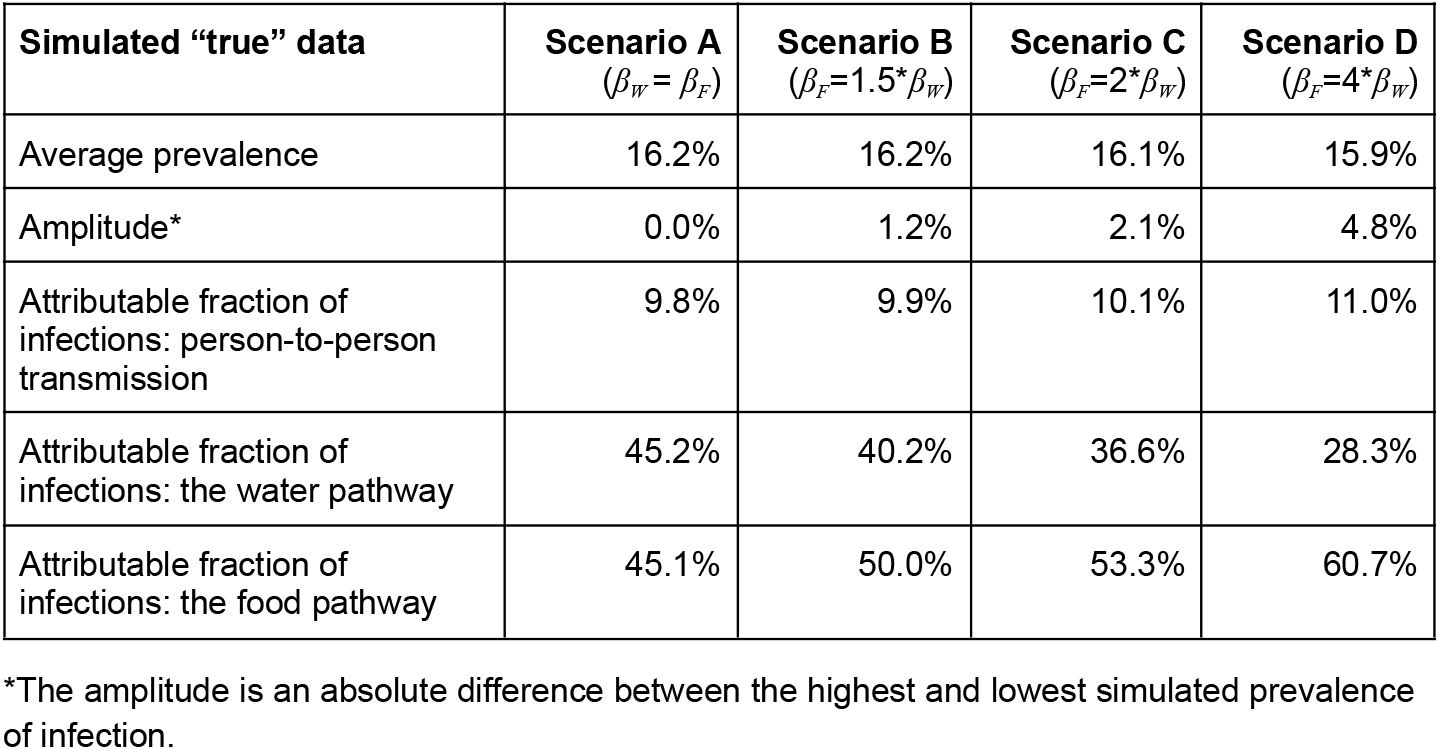
Simulated “true” prevalence of human infection and attributable fractions by transmission pathway (Scenario *iv* – *F(t)* and *W(t)* peak six months apart).

| **Simulated “true” data** | **Scenario A**<br>($\beta_W = \beta_F$) | **Scenario B**<br>($\beta_F = 1.5 * \beta_W$) | **Scenario C**<br>($\beta_F = 2 * \beta_W$) | **Scenario D**<br>($\beta_F = 4 * \beta_W$) |
| --- | --- | --- | --- | --- |
| Average prevalence | 16.2% | 16.2% | 16.1% | 15.9% |
| Amplitude* | 0.0% | 1.2% | 2.1% | 4.8% |
| Attributable fraction of infections: person-to-person transmission | 9.8% | 9.9% | 10.1% | 11.0% |
| Attributable fraction of infections: the water pathway | 45.2% | 40.2% | 36.6% | 28.3% |
| Attributable fraction of infections: the food pathway | 45.1% | 50.0% | 53.3% | 60.7% |

\*The amplitude is an absolute difference between the highest and lowest simulated prevalence of infection.

**Supplementary Table 5.** Estimated transmission rates for person-to-person, water-to-person, and food-to-person pathways across 16 scenarios using profile likelihood analysis.

| Transmission rates | True value | Estimated value (Chi-square 95% CIs) |  |  |  |
| --- | --- | --- | --- | --- | --- |
|  |  | Scenario *i*<br>(same seasonality) | Scenario *ii*<br>(one month apart) | Scenario *iii*<br>(three months apart) | Scenario *iv*<br>(opposite seasonality) |
| Scenario A |  |  |  |  |  |
| $\beta_I$ | 0.05 | 0 (0 - 0) | 0 (0 - 0) | 0 (0 - 0.002) | 0.465 (0.429 - 0.465) |
| $\beta_W$ | 0.075 | 0.06 (0 - 0.105) | 0.065 (0.063-0.067) | 0.074 (0.072-0.076) | 0.001 (0 - 0.009) |
| $\beta_F$ ($\beta_F=\beta_W$) | 0.075 | 0.004 (0 - 0.108) | 0.047 (0.044-0.048) | 0.068 (0.067-0.071) | 0 (0 - 0.075) |
| Scenario B |  |  |  |  |  |
| $\beta_I$ | 0.05 | 0 (0 - 0) | 0 (0 - 0) | 0 (0 - 0.001) | 0.42 (0.33 - 0.42) |
| $\beta_W$ | 0.06 | 0.06 (0 - 0.105) | 0.047 (0.046 - 0.049) | 0.062 (0.06 - 0.063) | 0 (0 - 0.016) |
| $\beta_F$ ($\beta_F=1.5*\beta_W$) | 0.09 | 0.047 (0 - 0.108) | 0.065 (0.063 - 0.068) | 0.081 (0.079 - 0.083) | 0.024 (0.020 - 0.041) |
| Scenario C |  |  |  |  |  |
| $\beta_I$ | 0.05 | 0 (0 - 0) | 0 (0 - 0) | 0 (0 - 0.002) | 0.36 (0.21 - 0.36) |
| $\beta_W$ | 0.05 | 0.06 (0 - 0.105) | 0.036 (0.034 - 0.038) | 0.05 (0.049 - 0.052) | 0 (0 - 0.022) |
| $\beta_F$ ($\beta_F=2*\beta_W$) | 0.1 | 0.004 (0 - 0.108) | 0.077 (0.074 - 0.080) | 0.095 (0.092 - 0.097) | 0.041 (0.038 - 0.067) |
| Scenario D |  |  |  |  |  |
| $\beta_I$ | 0.05 | 0 (0 - 0) | 0 (0 - 0) | 0 (0 - 0.001) | 0.09 (0.015 - 0.185) |
| $\beta_W$ | 0.03 | 0.06 (0 - 0.105) | 0.014 (0.012 - 0.016) | 0.025 (0.024 - 0.026) | 0.019 (0.010 - 0.029) |
| $\beta_F$ ($\beta_F=4*\beta_W$) | 0.12 | 0.047 (0 - 0.108) | 0.101 (0.098 - 0.104) | 0.125 (0.121 - 0.128) | 0.112 (0.098 - 0.129) |

Abbreviations: CI, confidence interval; NI, not identifiable;

**Supplementary Table 6.** Estimated transmission rates for person-to-person, water-to-person, and food-to-person pathways under the alternative scenario where *β*_*I*_ was predefined to be the largest.

| Transmission rates | True value | Estimated value (Chi-square 95% CIs) |  |  |  |
| --- | --- | --- | --- | --- | --- |
|  |  | Scenario *i*<br>(same seasonality) | Scenario *ii*<br>(one month apart) | Scenario *iii*<br>(three months apart) | Scenario *iv*<br>(opposite seasonality) |
| $\beta_I$ | 0.11 | 0 (0-0.002) | 0 (0-0.004) | 0.050<br>(0.027-0.073) | 0 (0-0.15) |
| $\beta_W$ | 0.03 | 0.054 (0-0.1) | 0.043<br>(0.039-0.046) | 0.034<br>(0.030-0.039) | 0.040<br>(0-0.042) |
| $\beta_F$ ($\beta_F=2*\beta_W$) | 0.06 | 0.056<br>(0-0.112) | 0.069<br>(0.064-0.074) | 0.065<br>(0.059-0.072) | 0.077<br>(0.03-0.082) |

## Supplementary Figures

**Supplementary Figure 1.**
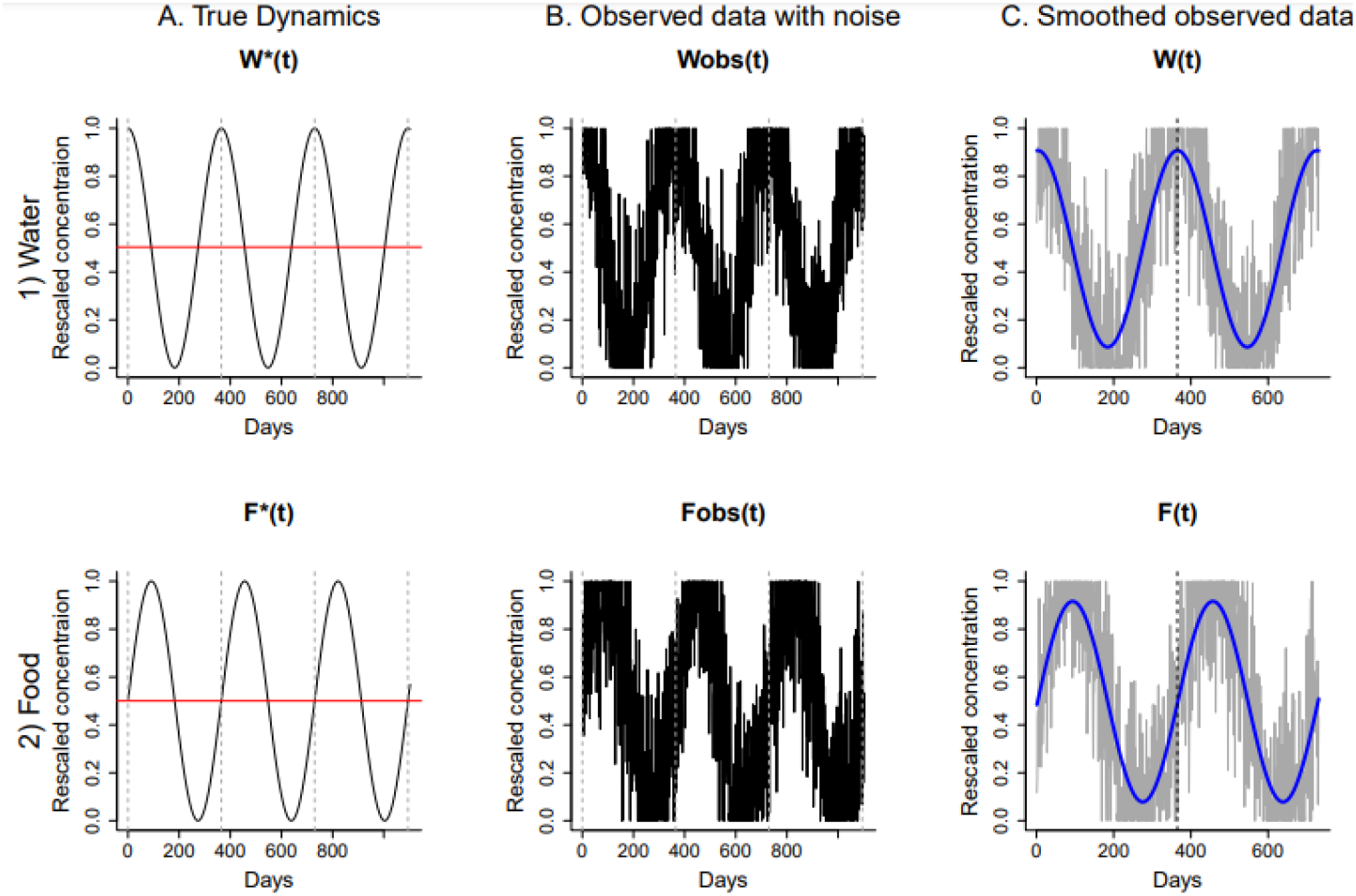
Simulated daily time series data on pathogen concentration in water and food (Scenario ii). This figure represents the simulated pathogen concentration data for Scenario ii in which the peak timings in water contamination and food contamination are three months apart, with water contamination peaking earlier. **Panel A** represents the true pathogen concentration simulated by Eq. 6-7. Vertical black dashed lines represent January 1, and horizontal red lines represent the mean of the rescaled concentration (which is set to 0.5). These true concentration data were used to simulate the true prevalence of human infection. **Panel B** represents the simulated daily time series for observed data, which was created by adding noise to the true concentration data in Panel A. **Panel C** represents the smoothed daily time series for observed data, which was created by fitting sine curves to the observed data in Panel B. These smoothed data were used to estimate transmission rates. (Note that the first year of data were removed.) The top row represents water contamination and the bottom row represents food contamination.

## References

1. Kotloff KL, Nataro JP, Blackwelder WC, Nasrin D, Farag TH, Panchalingam S, et al. Burden and aetiology of diarrhoeal disease in infants and young children in developing countries (the Global Enteric Multicenter Study, GEMS): a prospective, case-control study. The Lancet. 2013;382:209–222. doi:10.1016/S0140-6736(13)60844-2

2. The MAL-ED Network Investigators. The MAL-ED Study: A Multinational and Multidisciplinary Approach to Understand the Relationship Between Enteric Pathogens, Malnutrition, Gut Physiology, Physical Growth, Cognitive Development, and Immune Responses in Infants and Children Up to 2 Years of Age in Resource-Poor Environments. Clin Infect Dis. 2014;59:S193–S206. doi:10.1093/cid/ciu653

3. World Health Organization. WHO estimates of the global burden of foodborne diseases: foodborne disease burden epidemiology reference group 2007-2015. Geneva: World Health Organization; 2015. Available: https://apps.who.int/iris/handle/10665/350185

4. Pires SM, Evers EG, van Pelt W, Ayers T, Scallan E, Angulo FJ, et al. Attributing the human disease burden of foodborne infections to specific sources. Foodborne Pathog Dis. 2009;6:417–424. doi:10.1089/fpd.2008.0208

5. Evers EG, Fels-Klerx HJ van der, Nauta MJ, Schijven JF, Havelaar AH. Campylobacter source attribution exposure assessment. Int J Risk Assess Manag. 2008;8:174–190. doi:10.1504/IJRAM.2008.016151

6. Shioda K, Brouwer AF, Lamar F, Mucache HN, Levy K, Freeman MC. Opportunities to Interrupt Transmission of Enteropathogens of Poultry Origin in Maputo, Mozambique: A Transmission Model Analysis. Environ Health Perspect. 131: 117004. doi:10.1289/EHP12314

7. Brouwer AF, Eisenberg MC, Bakker KM, Boerger SN, Zahid MH, Freeman MC, et al. Leveraging infectious disease models to interpret randomized controlled trials: controlling enteric pathogen transmission through water, sanitation, and hygiene interventions. Epidemiology; 2022 Apr. doi:10.1101/2022.04.28.22274441

8. Kraay ANM, Hayashi MAL, Hernandez-Ceron N, Spicknall IH, Eisenberg MC, Meza R, et al. Fomite-mediated transmission as a sufficient pathway: a comparative analysis across three viral pathogens. BMC Infect Dis. 2018;18:540. doi:10.1186/s12879-018-3425-x

9. Caldwell JM, LaBeaud AD, Lambin EF, Stewart-Ibarra AM, Ndenga BA, Mutuku FM, et al. Climate predicts geographic and temporal variation in mosquito-borne disease dynamics on two continents. Nat Commun. 2021;12:1233. doi:10.1038/s41467-021-21496-7

10. Stevens EL, Carleton HA, Beal J, Tillman GE, Lindsey RL, Lauer AC, et al. Use of Whole Genome Sequencing by the Federal Interagency Collaboration for Genomics for Food and Feed Safety in the United States. J Food Prot. 2022;85:755–772. doi:10.4315/JFP-21-437

11. Vogel M, Utpatel C, Corbett C, Kohl TA, Iskakova A, Ahmedov S, et al. Implementation of whole genome sequencing for tuberculosis diagnostics in a low-middle income, high MDR-TB burden country. Sci Rep. 2021;11:15333. doi:10.1038/s41598-021-94297-z

12. World Health Organization. Whole genome sequencing for foodborne disease surveillance: landscape paper. 2018. Available: https://iris.who.int/bitstream/handle/10665/272430/9789241513869-eng.pdf

13. Rothstein AP, Jesser KJ, Feistel DJ, Konstantinidis KT, Trueba G, Levy K. Population genomics of diarrheagenic Escherichia coli uncovers high connectivity between urban and rural communities in Ecuador. Infect Genet Evol. 2023;113:105476. doi:10.1016/j.meegid.2023.105476

14. Wang Y, Mairinger W, Raj SJ, Yakubu H, Siesel C, Green J, et al. Quantitative assessment of exposure to fecal contamination in urban environment across nine cities in low-income and lower-middle-income countries and a city in the United States. Sci Total Environ. 2022;806:151273. doi:10.1016/j.scitotenv.2021.151273

15. Moe CL, Sobsey MD, Samsa GP, Mesolo V. Bacterial indicators of risk of diarrhoeal disease from drinking-water in the Philippines. Bull World Health Organ. 1991;69:305–317.

16. McMurry TL, McQuade ETR, Liu J, Kang G, Kosek MN, Lima AAM, et al. Duration of Postdiarrheal Enteric Pathogen Carriage in Young Children in Low-resource Settings. Clin Infect Dis. 2021;72:e806–e814. doi:10.1093/cid/ciaa1528

17. Tribble DR, Baqar S, Scott DA, Oplinger ML, Trespalacios F, Rollins D, et al. Assessment of the Duration of Protection in Campylobacter jejuni Experimental Infection in Humans. Infect Immun. 2010;78:1750–1759. doi:10.1128/IAI.01021-09

18. Missouri Depertment of Health and Senior Services. Campylobacteriosis. [cited 13 Jan 2025]. Available: https://health.mo.gov/safety/foodsafety/foodborneillness/campylobacteriosis.php

19. Cawthraw SA, Feldman RA, Sayers AR, Newell DG. Long-term antibody responses following human infection with Campylobacter jejuni. Clin Exp Immunol. 2002;130:101–106. doi:10.1046/j.1365-2249.2002.01966.x

20. Wang H, Abbas KM, Abbasifard M, Abbasi-Kangevari M, Abbastabar H, Abd-Allah F, et al. Global age-sex-specific fertility, mortality, healthy life expectancy (HALE), and population estimates in 204 countries and territories, 1950–2019:a comprehensive demographic analysis for the Global Burden of Disease Study 2019. The Lancet. 2020;396:1160–1203. doi:10.1016/S0140-6736(20)30977-6

21. Knee J, Sumner T, Adriano Z, Berendes D, Bruijn E de, Schmidt W-P, et al. Risk factors for childhood enteric infection in urban Maputo, Mozambique: A cross-sectional study. PLoS Negl Trop Dis. 2018;12:e0006956. doi:10.1371/journal.pntd.0006956

22. Raue A, Kreutz C, Maiwald T, Bachmann J, Schilling M, Klingmüller U, et al. Structural and practical identifiability analysis of partially observed dynamical models by exploiting the profile likelihood. Bioinformatics. 2009;25:1923–1929. doi:10.1093/bioinformatics/btp358

23. Murphy SA, Van Der Vaart AW. On Profile Likelihood. J Am Stat Assoc. 2000;95:449–465. doi:10.1080/01621459.2000.10474219

24. Venzon DJ, Moolgavkar SH. A Method for Computing Profile-Likelihood-Based Confidence Intervals. Appl Stat. 1988;37:87. doi:10.2307/2347496

25. Hubbard, Sydney C. Evaluating the efficacy of water, sanitation, and hygiene interventions in the context of climate change with a focus on urban, coastal Mozambique. ProQuest Dissertations & Theses; 2023. Available: https://search.libraries.emory.edu/catalog/9937789011402486

26. Nguyen TK, Bui HT, Truong TA, Lam DN, Ikeuchi S, Ly LKT, et al. Retail fresh vegetables as a potential source of Salmonella infection in the Mekong Delta, Vietnam. Int J Food Microbiol. 2021;341:109049. doi:10.1016/j.ijfoodmicro.2021.109049

27. Schwan CL, Desiree K, Bello NM, Bastos L, Hok L, Phebus RK, et al. Prevalence of Salmonella enterica Isolated from Food Contact and Nonfood Contact Surfaces in Cambodian Informal Markets. J Food Prot. 2021;84:73–79. doi:10.4315/JFP-20-112

28. Chua PLC, Huber V, Ng CFS, Seposo XT, Madaniyazi L, Hales S, et al. Global projections of temperature-attributable mortality due to enteric infections: a modelling study. Lancet Planet Health. 2021;5:e436–e445. doi:10.1016/S2542-5196(21)00152-2

